# Recording sampling rate as a negative-control exposure for detecting measurement-driven bias in heart-rate-variability–outcome associations: a quantitative bias analysis across ICU waveform databases

**DOI:** 10.64898/2026.11.15.26368154

**Authors:** Hui Ye, Qian Zhang, Song Chen, Peiwu Li, Jian Wan, Yilong Hao

## Abstract

**Importance:** Heart-rate variability (HRV) is studied extensively as a prognostic marker in critical illness, yet the electrocardiographic sampling rate at which HRV is computed systematically distorts the resulting values. Whether the associations between HRV and clinical outcomes reported in the literature are partly a consequence of this distortion has not been examined directly.

**Objective:** To evaluate whether between-database differences in native ECG sampling rate can be used as a negative-control exposure to detect measurement-driven bias in HRV–outcome associations, and to quantify the sensitivity of a published association to an empirically measured distortion coefficient using quantitative bias analysis (QBA).

**Design:** A methods investigation in three parts. Part I sets out the causal structure and states why a strict instrumental-variable (IV) design is inappropriate here. Part II derives a bias-correction factor under a multiplicative measurement-error model and propagates the empirical confidence interval of the distortion coefficient as a sensitivity band. Part III applies the procedure to an observed HRV–mortality association from a single ICU waveform database, and examines the separate bias arising from threshold-based categorisation.

**Setting and data sources:** Two critical-care waveform databases with different native sampling rates (MIMIC-IV Waveform, approximately 250 Hz; MIMIC-III Waveform Matched p04, 125 Hz) and an independent 500-Hz reference cohort (PTB-XL) for mechanism validation. The outcome association was estimated in 1,445 ICU stays from MIMIC-III p04, of whom 141 died in hospital (9.8%).

**Exposures:** Native ECG sampling rate as a negative-control exposure; SDNN, RMSSD and heart rate as the measured exposures whose associations with mortality are evaluated.

**Main outcomes and measures:** In-hospital mortality. Distortion coefficients were obtained by end-to-end reprocessing of 7,293 original waveform segments with a fixed QRS detector. Corrected odds ratios were reported as QBA sensitivity bands rather than point-identified effects. Misclassification was quantified as the proportion of recordings below a 20 ms HRV threshold reclassified above it by coarse sampling.

**Results:** End-to-end reprocessing gave an RMSSD distortion coefficient of 1.1575 (95% CI 1.0835–1.2492; 2,276 segments in 188 patients) for 250→62.5 Hz and 1.0588 (95% CI 1.0294–1.1033; 4,988 segments) for 125→62.5 Hz; high-frequency power was more affected than low-frequency power (1.2033 vs 1.0716). Under the multiplicative model, an observed crude RMSSD–mortality odds ratio of 1.802 corresponded to a corrected interval of 1.8335–1.915 across the coefficient’s 95% CI. The correction was small relative to confounding: the same association fell to 1.116 after age adjustment. Coarse sampling reclassified 33.70% of low-HRV recordings in MIMIC-IV and 20.72% in MIMIC-III, a larger potential source of bias than the continuous-scale correction. The cross-database negative-control gradient was completed after linking both waveform cohorts to hospital outcomes: RMSSD inflation was reproducible (median ×1.101 at 125→62.5 Hz in MIMIC-III and ×1.270 at 250→62.5 Hz in MIMIC-IV), but the age-adjusted HRV–mortality risk ratio did not increase at lower rates (MIMIC-III 1.70→1.21; MIMIC-IV 1.31→1.29→1.20; all 95% CIs crossed 1). The falsification test was therefore negative in these cohorts, and we report it as such rather than as evidence that measurement bias is absent.

**Conclusions and Relevance:** Sampling rate is a detectable and correctable source of bias in HRV–outcome research. Recording the sampling rate of each signal, stratifying effects by baseline variability, and reporting a QBA sensitivity band before dichotomising HRV would make published associations more interpretable. The object of inference here is the measurement-error method, not a new clinical biomarker; the proposed reporting steps require no additional data collection.

## 1. Introduction

Reduced heart-rate variability (HRV) is one of the most widely reported physiological correlates of critical illness. Reviews in critical care describe inverse associations between HRV and severity or mortality across diagnostic groups [6,9,10], and cohort studies have reported that changes in HRV precede the clinical recognition of sepsis [7,8]. These findings have encouraged proposals that HRV might serve as a prognostic biomarker at the bedside [6]. The literature is also heterogeneous. Sampling protocols, segment lengths, QRS detectors and frequency-band definitions differ substantially between studies, and reviews have noted that part of the apparent HRV–mortality association tracks heart rate rather than autonomic variability [9,10].

A separate literature concerns how digital sampling affects HRV measurement. Hejjel and Roth, using model tachograms, showed that time-domain error rises steeply as the sampling interval widens: at a 10 ms interval, the relative accuracy error for SDNN exceeded 30% in low-variability series [2]. Burma and colleagues downsampled 1,000-Hz ECG recordings and found that RMSSD required at least 90 Hz, and SDNN at least 60 Hz, to agree with the reference recording [3]. Zhou and colleagues recently derived analytical error bounds for HR, RMSSD and SDNN and validated them in 58 volunteers, providing quantitative guidance for sampling-rate selection [1]. Other work has examined interpolation and detector choice [4,5]. The magnitude of the distortion also depends on how downsampling is performed: a recent preprint showed that extrema-preserving schemes retain QRS-detection accuracy and RMSSD down to 12 Hz in the MIT-BIH databases, whereas naïve decimation fails below 45 Hz [43]. Our reanalysis uses straight decimation, as in the original pipeline, so the coefficients reported here describe that specific processing choice rather than sampling rate alone. What these studies share is a focus on measurement accuracy. They stop at the question of how far a computed HRV value departs from its true value.

The two literatures have not been brought together. Although reviewers of critical-care HRV have noted sampling-rate heterogeneity as a source of between-study inconsistency [8,10], we are aware of no study that treats the between-database difference in native sampling rate as a diagnostic instrument for bias in an HRV– outcome association, or that uses a measured distortion coefficient to correct such an association.

Here we make three claims. First, sampling rate can be used as a negative-control exposure in the sense used by Lipsitch and colleagues and elaborated by Arnold and colleagues: a variable that should not be related to the outcome through the biological pathway of interest, but that shares the measurement and selection processes capable of generating a spurious association [20,21]. Second, because the distortion coefficient can be estimated directly from original waveforms, the magnitude of the resulting bias can be propagated formally rather than merely acknowledged, following the logic of quantitative bias analysis [11,13,14,18]. Third, the categorisation of HRV at a fixed threshold introduces a distinct misclassification problem that may dominate the continuous-scale correction.

We also state one claim we do *not* make. We do not propose sampling rate as a valid instrumental variable. Recording rate is correlated with centre, device era and monitoring intensity, so both independence from confounders and the exclusion restriction fail. We therefore do not attempt causal identification. The design is a bias audit: we ask how sensitive an observed association is to a measured measurement-error process, and we report a sensitivity band rather than a corrected point estimate with a claim of validity.

The manuscript is organised in three parts. Part I defines the causal structure and the negative-control logic. Part II derives the correction factor and specifies how the distortion coefficient is propagated. Part III applies the procedure to a real outcome association and examines threshold misclassification. The same distortion coefficients are used throughout, and they were estimated from the waveforms before any outcome association was considered; this separation is the methodological discipline of the design.

## 2. Methods

### 2.1 Causal structure and the negative-control exposure

Let *S* denote the native sampling rate of the ECG used to derive an RR-interval series, *H\** the underlying physiological HRV signal, *H*obs the value computed from the digitised signal, *C* a set of common causes (age, illness severity, centre, device era, monitoring intensity, mean heart rate), and *Y* the clinical outcome. Sampling rate affects the observed value through R-peak time quantisation, *S* → *H*obs → *Y*; the physiological signal enters the observation through *H\** → *H*obs; and the common causes influence *S, H*obs and *Y*.

The negative-control premise is that *S* has no pathway to *Y* other than through the measurement of *H\** and through its association with *C*. If the *H*obs–*Y* association is entirely physiological, conditioning on *C* should remove any dependence on *S*. An association that persists, and whose gradient across sampling-rate tiers matches the direction and magnitude of the independently measured distortion coefficients, is evidence that measurement artifact contributes to the estimate. Negative controls of this form are best understood as a falsification test, not as a proof: as Arnold and colleagues emphasise, they can indicate the presence of bias without determining its direction or magnitude [21], and a null result cannot establish that bias is absent [20,22]; methods that exploit negative controls for partial identification, such as the control-outcome calibration approach [23], require assumptions we are not in a position to make here.

The choice rules out a strict IV analysis. For *S* to be a valid instrument it would need to be independent of *C*, which is contradicted by the clustering of recording equipment within centres and device generations, and it would need to affect *Y* only through the measured HRV, which is contradicted by the fact that sampling rate is a marker of the surrounding monitoring environment. Causal diagrams for measurement bias [28,29] and for monitoring-driven confounding in intensive care [30] make the same point from different directions: the monitoring process is itself related to patient state. We therefore use *S* only as a bias scale, and we interpret the resulting estimates as sensitivity analyses in the sense of Greenland and of Lash and colleagues [11,14].

### 2.2 Estimation of the distortion coefficients

The distortion coefficients were estimated by end-to-end reprocessing of original waveforms, using the same detection and HRV pipeline at each sampling rate. A fixed QRS detector (wfdb `gqrs`) was applied to 7,293 segments: 2,294 from MIMIC-IV at native 250 Hz and 4,999 from MIMIC-III p04 at native 125 Hz. Segments were then downsampled in stages to 62.5 Hz, and HRV indices were recomputed from the detected beat times at each stage. All HRV values used in this analysis were recomputed in this way; none were read from the intermediate tables used for the earlier submission. The procedure, its known limitations (45 segments without a standard ECG lead fell back to the first available channel; a stated ≥60 s exclusion was not applied in the original pipeline), and full statistics are documented in the companion direction-1 reanalysis and in the archived code and aggregates [41].

The primary coefficient for RMSSD was 1.1575 (95% CI 1.0835–1.2492; 2,276 segments in 188 patients) for 250→62.5 Hz, and 1.0588 (95% CI 1.0294–1.1033; 4,988 segments) for 125→62.5 Hz. Low-frequency power increased by a factor of 1.0716 and high-frequency power by 1.2033 in the MIMIC-IV material (1,405 valid segments); in MIMIC-III the corresponding high-frequency coefficient was 1.0913 (3,103 segments). Because HF is more affected than LF, the LF/HF ratio is not rate-invariant, a point that follows directly from band-resolved measurement error rather than from any physiological claim.

As a check on the mechanism, an independent 500-Hz reference cohort (PTB-XL, 21,755 recordings from 18,844 patients [34]) reproduced the dose–response (1.0039, 1.0224 and 1.1217 at 250, 125 and 62.5 Hz). Replacing crude integer R-peak timing with parabolic sub-sample peak refinement reduced the 62.5-Hz ratio from 1.1217 to 1.0195, consistent with timing quantisation rather than anti-alias filtering as the dominant mechanism.

The analytic dataset used for the outcome association was the MIMIC-III p04 cohort [32,33]: 1,445 ICU stays, of which 141 ended in hospital death (9.8%), with HRV features extracted from 40,595 successfully processed segments (24,518 segments of at least five minutes in 1,064 subjects). The sampling rate of this database is 125 Hz, which is why the 1.0588 coefficient is the relevant one for the main correction. MIMIC-III, MIMIC-IV and PTB-XL are cited according to their PhysioNet records [31–35], and HRV computation follows the Task Force conventions where applicable [36].

### 2.3 Quantitative bias analysis under multiplicative error

Suppose the measured exposure is a multiplicative distortion of the physiological one, *H*obs = δ *H\**. Under a logistic model in the true exposure, logit Pr(*Y*) = β□ + β *H\**, fitting against *H*obs yields a slope of approximately β/δ when the error is non-differential and the model is correctly specified. On the odds-ratio scale this gives

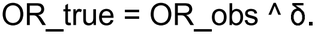

The derivation assumes a constant multiplicative factor over the exposure range; in practice δ varies with baseline variability and frequency band, so the result should be read as a first-order approximation. The direction of the adjustment is worth stating explicitly. Because δ > 1 inflates the exposure scale, the slope estimated against the inflated values is compressed towards the null; the correction therefore moves the odds ratio *away* from 1 on the multiplicative scale. That the correction increases an already positive odds ratio is not evidence that the association is an artifact; it is a consequence of the error model, and it is why the size of the correction must be compared with the size of the association and with changes under confounding adjustment.

Following the good-practice recommendations for QBA [12,13], we treat δ as a bias parameter with an empirical distribution rather than a fixed constant. The 95% confidence interval of the measured coefficient is used as the sensitivity band, giving OR_true(δ_lo) to OR_true(δ_hi). This is a sensitivity analysis, not probabilistic bias analysis: we do not assign a probability distribution to δ, and we do not report a single “corrected” estimate as if it were identified. No external validation data were needed because the bias parameter came from the end-to-end waveform reprocessing described above [11,18].

For SDNN, which has no separately estimated coefficient, the RMSSD time-domain coefficient is borrowed and the approximation is flagged in the results. Heart rate is retained as a negative control exposure in the strict sense: its measurement depends on R-R interval counting and should not be distorted by the sampling rates considered here, so δ = 1 and no correction is applied. If sampling rate were generating associations indiscriminately, the heart-rate association would move as well; it does not.

### 2.4 Threshold misclassification

Separate from the continuous-scale correction, we examined the effect of coarse sampling on a common reporting practice: classifying HRV as low when it falls below a 20 ms threshold. Using the reprocessed waveforms, we computed the proportion of recordings below the threshold at the reference rate that were moved above it by downsampling. This is an outcome-independent misclassification of the exposure, and its consequences for an odds ratio depend on the prevalence of low HRV and on the true association; we therefore report the reclassification fractions as the primary quantity and treat their effect on association estimates as a sensitivity consideration rather than a precise correction [18,38,39,40].

### 2.5 What this analysis does not do

We report the cross-database gradient using the waveform subsets that could be linked to hospital outcomes, and we report it as a negative result: the linked cohorts were small (10 and 35 deaths), so the test is descriptive rather than confirmatory. Two further omissions are deliberate. We do not construct a negative-control outcome, because a candidate would need to share the confounders of HRV without lying on the autonomic pathway to mortality; none could be constructed credibly from the available tables, and inventing one would defeat the purpose of the test [20,21]. We do not present a clinical recommendation: the cohort association used here is counter-intuitive in direction and collapses under age adjustment, a finding already documented in the source report, and it is used because it is a demanding test case for the method, not because it is a candidate biomarker.

Our approach is complementary to recent work that combines negative controls with empirical calibration to attribute a share of an observed association to systematic bias [19,26,27]. That work targets residual confounding and selection processes in treatment-effect studies and reports a bias fraction on a bounded scale. A related line uses panels of negative-control *outcomes* to diagnose and calibrate shared systematic error in real-world evidence [42]: the negative controls there are clinical endpoints chosen to share the confounders of the exposure, whereas ours is a physical property of the signal that has no clinical pathway to mortality. Ours targets a specified physical measurement process with an externally measured bias parameter and reports a corrected interval. External adjustment [16] and bounding approaches [17] likewise require an assumed magnitude for a confounder and return an adjusted estimate; the multiple-bias framework of Greenland [15] addresses several such sources jointly. It is also distinct from methods for informative monitoring in longitudinal EHR data [30]: those methods address the timing and frequency of clinical measurements as time-varying confounding, whereas we address the sampling rate of the ECG signal itself.

Finally, the E-value [24,25] answers a different question — how strong an unmeasured confounder would need to be — and is not a substitute for a bias parameter measured from the data.

### 2.6 Reproducibility

All corrections were produced by the archived analysis script, which reads the observed odds ratios and distortion coefficients embedded in the script (both transcribed from the local sources listed above), computes OR_true = OR_obs^δ at the point estimate and interval endpoints, and writes `qba_results.csv` and `qba_report.txt`. The figure is generated from the same table. Distortion coefficients and code are archived in Zenodo v2 [41]; under the PhysioNet data-use agreement, neither waveforms nor per-segment tables are deposited [35].

## 3. Results

### 3.1 Distortion coefficients

Table 1 summarises the measured coefficients. RMSSD inflation ranged from 1.0588 at 125→62.5 Hz to 1.1575 at 250→62.5 Hz. The frequency-domain contrast was larger: HF power increased by 1.2033 in MIMIC-IV against 1.0716 for LF over the same downsampling step. Heart rate, used as the negative-control exposure, had a coefficient of 1.0000 by construction of the detection method.

**Table 1.** Distortion coefficients measured by end-to-end reprocessing of original waveforms. Ratios are relative to the reference sampling rate in each row; all values recomputed with a fixed QRS detector (wfdb gqrs). CI, confidence interval.

| Index | Database | Downsampling step | Distortion coefficient δ (95% CI) | Segments |
|---|---|---|---|---|
| RMSSD | MIMIC-IV | 250 → 62.5 Hz | 1.1575 (1.0835–1.2492) | 2,276 (188 patients) |
| RMSSD | MIMIC-III p04 | 125 → 62.5 Hz | 1.0588 (1.0294–1.1033) | 4,988 |
| LF power | MIMIC-IV | 250 → 62.5 Hz | 1.0716 | 1,405 |
| HF power | MIMIC-IV | 250 → 62.5 Hz | 1.2033 | 1,405 |
| HF power | MIMIC-III p04 | 125 → 62.5 Hz | 1.0913 | 3,103 |
| Heart rate | both | any | 1.0000 (no distortion) | — |

### 3.2 Corrected association estimates

Table 2 gives the observed and corrected odds ratios. For the crude RMSSD–mortality association (OR_obs = 1.802), QBA gave OR_true = 1.8655 at δ = 1.0588, with a sensitivity band of 1.8335–1.915 over δ = 1.0294–1.1033. For SDNN the corresponding interval was 1.4181–1.4541 around a point estimate of 1.4323, using the borrowed time-domain coefficient. After age adjustment, the RMSSD association was 1.116 and the corrected interval was 1.1196–1.1287; under the stricter g-computation adjustment it was 1.154, corrected to 1.1589–1.1712. Heart rate was unchanged at 1.350, 1.609 and 1.451 in the three models.

**Table 2.** Observed and QBA-corrected odds ratios for in-hospital mortality. OR_obs from the MIMIC-III p04 cohort (n=1,445; 141 deaths). OR_true computed as OR_obs^δ; interval endpoints use the 95% CI of δ (1.0294–1.1033). “Borrowed δ” indicates the RMSSD time-domain coefficient used for SDNN. HR is shown as the rate-invariant control exposure (δ=1).

| Feature | Adjustment | OR_obs | δ (95% CI) | OR_true point | OR_true sensitivity band |
|---|---|---|---|---|---|
| SDNN | crude | 1.404 | 1.0588 (1.0294–1.1033), borrowed | 1.4323 | 1.4181–1.4541 |
| SDNN | age-adjusted | 1.018 | borrowed | 1.0191 | 1.0185–1.0199 |
| SDNN | strict g-computation | 1.148 | borrowed | 1.1574 | 1.1527–1.1645 |
| RMSSD | crude | 1.802 | 1.0588 (1.0294–1.1033) | 1.8655 | 1.8335–1.9150 |
| RMSSD | age-adjusted | 1.116 | 1.0588 (1.0294–1.1033) | 1.1232 | 1.1196–1.1287 |
| RMSSD | strict g-computation | 1.154 | 1.0588 (1.0294–1.1033) | 1.1638 | 1.1589–1.1712 |
| Heart rate | crude | 1.350 | 1.0000 | 1.3500 | 1.3500–1.3500 |
| Heart rate | age-adjusted | 1.609 | 1.0000 | 1.6090 | 1.6090–1.6090 |
| Heart rate | strict g-computation | 1.451 | 1.0000 | 1.4510 | 1.4510–1.4510 |

The multiplicative correction scaled with the magnitude of the observed association: the proportional shift was about 3.5% at the crude RMSSD estimate and under 1% at the age-adjusted estimate. In other words, the measurement correction is largest where the crude association is largest, and it becomes negligible once the association has been reduced by confounding adjustment. This ordering matters for interpretation: in these data, adjustment for measured confounders changed the estimate far more than the sampling-rate correction did.

### 3.3 Threshold misclassification

Coarse sampling moved 33.70% of MIMIC-IV recordings initially below 20 ms above the threshold (182 of 540 segments), and 20.72% of MIMIC-III recordings (246 of 1,187). Because a threshold defines the exposed group in many published HRV studies, this form of bias is not equivalent to the continuous-scale correction and can be larger in its effect on a reported odds ratio. Reporting the sampling rate of every signal is therefore a prerequisite for comparing studies that use the same nominal threshold.

### 3.4 Negative-control gradient

The direction required by the negative-control test is available from the distortion coefficients: lower sampling rates and lower baseline variability produce larger inflation, and HF is affected more than LF. Having linked both waveform cohorts to hospital outcomes, we estimated the outcome association within each sampling-rate tier. End-to-end segments were restricted to durations of at least 60 s, aggregated to the median RMSSD per subject per tier, and split at the tertiles; the age-adjusted risk ratio for high versus low variability was obtained by marginal standardisation from a logistic model, with 95% confidence intervals from 1,500 subject-level bootstrap resamples. The measurement gradient was reproducible — median RMSSD rose by ×1.101 from 125 to 62.5 Hz in MIMIC-III and by ×1.270 from 250 to 62.5 Hz in MIMIC-IV — but the association gradient was not: the risk ratio fell from 1.70 to 1.21 in MIMIC-III and from 1.31 to 1.20 in MIMIC-IV, and every interval crossed unity (Table 3, Figure 2). We report this as a negative falsification test, not as evidence that measurement bias is absent.

**Table 3.** Negative-control gradient: measurement scale and outcome association. The diagnostic requires both a measured bias scale and an observed association gradient; both are now reported.

| Requirement | Status | Source / result |
|---|---|---|
| Distortion increases as sampling rate falls (RMSSD 1.0588 at 125→62.5 vs 1.1575 at 250→62.5) | Available | End-to-end reanalysis [41], Table 1 |
| HF inflated more than LF (1.2033 vs 1.0716) | Available | End-to-end reanalysis [41], Table 1 |
| Rate-invariant control exposure (heart rate δ=1) | Available | Table 2 |
| RMSSD median inflation reproduced in linked cohorts (MIMIC-III ×1.101; MIMIC-IV ×1.270) | Completed | End-to-end segments linked to hospital outcomes; 97/163 subjects |
| HRV–mortality association by sampling-rate tier (age-adjusted RR) | **Completed — negative** | MIMIC-III 1.70→1.21; MIMIC-IV 1.31→1.29→1.20; all 95% CIs cross 1; 10/35 deaths |
| Negative-control outcome sharing confounders but not the autonomic pathway | **Not constructed** | No credible local instance; single-arm exposure test reported instead |

**Figure 1.**
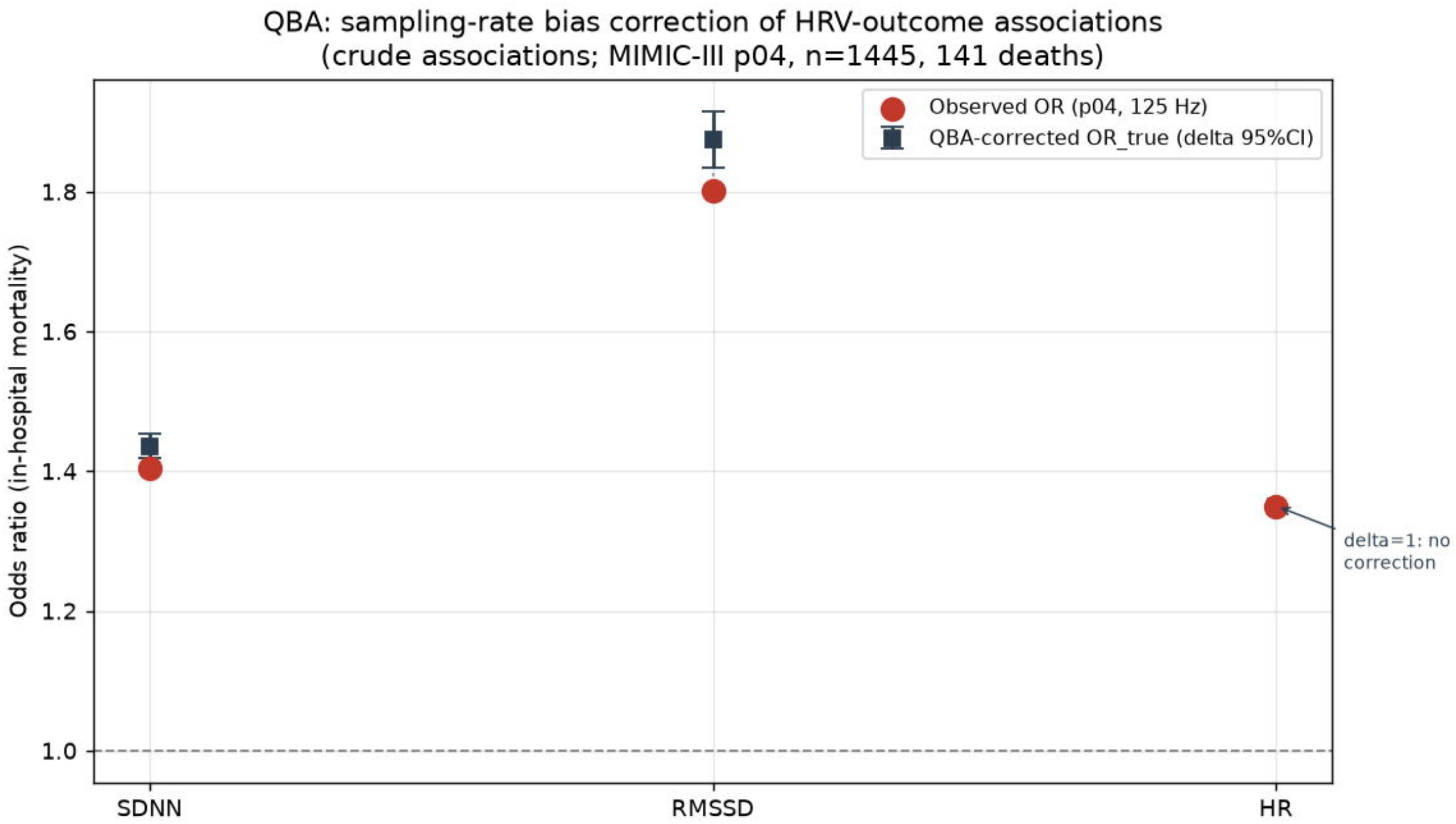
Observed versus QBA-corrected odds ratios. Observed odds ratios (filled markers) and corrected point estimates with sensitivity bands (open markers) for SDNN, RMSSD and heart rate across three adjustment models. Heart rate has δ=1 and therefore no correction.

**Figure 2.**
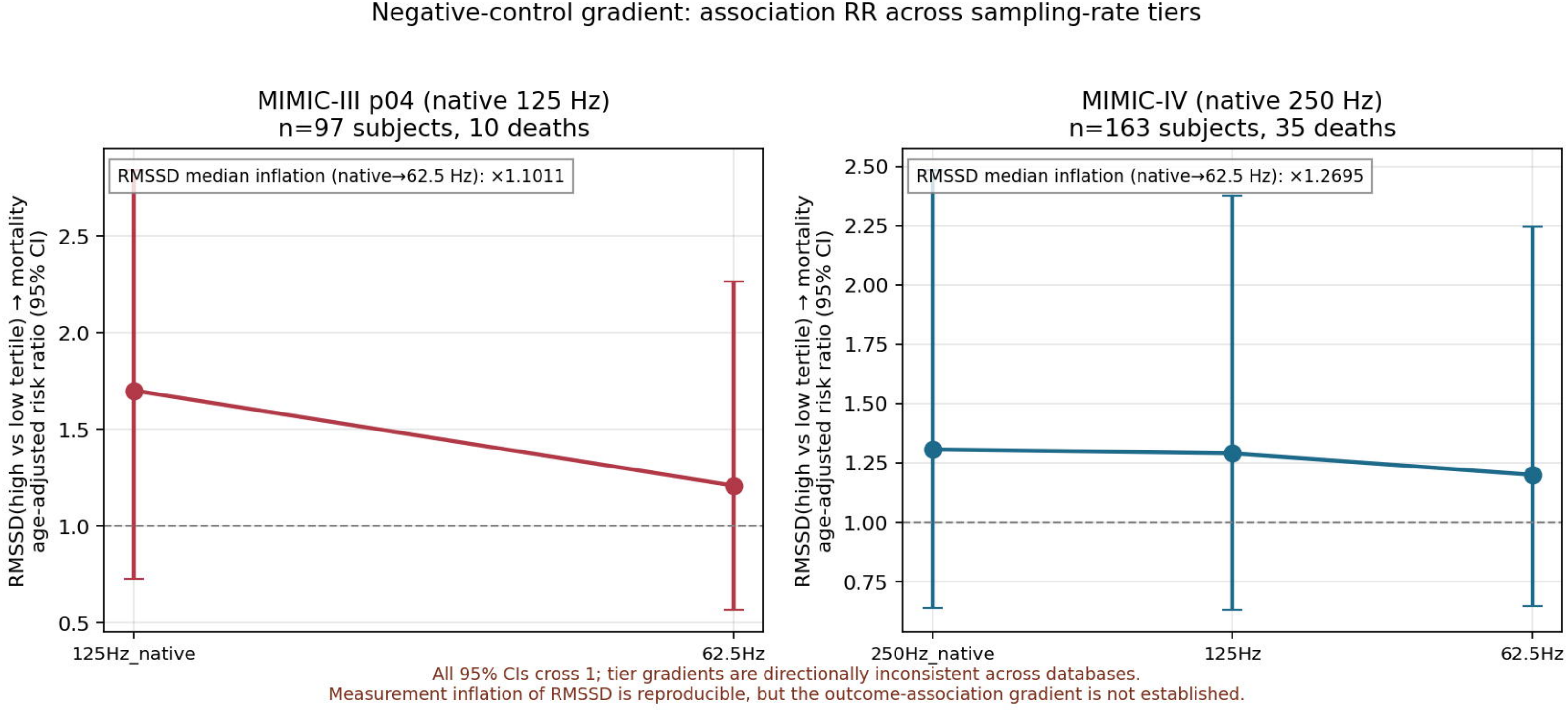
Negative-control gradient across sampling-rate tiers. Age-adjusted risk ratios for high versus low RMSSD (tertiles) with 95% bootstrap confidence intervals at each sampling-rate tier, in MIMIC-III p04 (left) and MIMIC-IV (right). The annotated values are subject-level median RMSSD inflation from the native rate to 62.5 Hz. All intervals cross unity and the association does not increase at lower rates; the panel is therefore a negative falsification test.

## 4. Discussion

We asked whether a known measurement artifact could account for part of the HRV– outcome associations that appear in the critical-care literature. Using sampling rate as a negative-control exposure and a measured distortion coefficient as a bias parameter, we found that the correction to a crude RMSSD–mortality odds ratio was small (1.802 to an interval of 1.8335–1.915), that it shrank to under 1% after confounder adjustment, and that threshold-based classification was a more substantial source of potential bias, moving around one fifth to one third of low-HRV recordings across the threshold. The results do not show that published HRV associations are artifacts. They show that the artifact has a measurable, bounded effect, and that the effect can be reported rather than ignored.

The negative-control framing is useful precisely because it separates two hypotheses. A genuinely physiological HRV–outcome association should not depend on the recording equipment’s sampling rate. An association that strengthens at lower rates, and by an amount compatible with δ, would implicate measurement. With the outcome linkage now in place, the falsification test could be run in full. It returned a negative result: although the measurement inflation was reproducible, the HRV–mortality association did not strengthen at lower sampling rates in either cohort, and none of the tier-specific risk ratios excluded unity. Read as a falsification test, this does not confirm a measurement-driven association; nor, with 10 and 35 deaths in the two cohorts, does it exclude one. The defensible reading is that the sampling-rate gradient is not a sensitive diagnostic at the sample sizes available here, which is precisely why the QBA sensitivity band — rather than the gradient — carries the quantitative argument.

Several features of the design merit emphasis. The bias parameter was estimated from original waveforms rather than assumed, which is the condition under which QBA is most informative [11,18,19]. Negative-control and QBA approaches were used as diagnostics and sensitivity tools rather than as instruments for identification [20–23]. Heart rate, which should be rate-invariant under the detection method, was carried through every model as a control exposure and remained unchanged. And the two bias mechanisms — continuous inflation and threshold misclassification — were quantified separately, because a single correction would obscure the larger of the two.

The limitations are substantial. Sampling rate is collinear with centre and device generation in these databases, so the negative-control exposure is not independent of the care environment; this is the reason for avoiding an IV interpretation and for presenting a sensitivity band rather than a causal effect. The benchmark HRV–outcome association used for the QBA comes from one database at one native rate. The cross-database gradient that we were able to run is underpowered: the linked waveform subsets contained 97 subjects with 10 deaths in MIMIC-III and 163 subjects with 35 deaths in MIMIC-IV, and the tier-specific confidence intervals were wide and all crossed unity. Sampling rate is also strongly confounded with centre and device generation in these databases, and the sampling-rate tiers within a cohort arise from reprocessing the same recordings rather than from independent devices, so the gradient is a within-recording sensitivity analysis and not a between-device comparison. The exposure units of the source odds ratios are not documented in enough detail in the source report to make the correction more precise than a first-order approximation, and the SDNN coefficient is borrowed from RMSSD. PTB-XL segments are ten seconds long and cannot support LF/HF analysis, so frequency-domain validation rests on the ICU waveforms alone. Finally, the source association is counter-intuitive in direction and attenuates sharply with age adjustment; we use it as a test case for the method, not as evidence for a clinical claim.

These constraints point to a modest reporting recommendation rather than a new analytical standard. Studies reporting HRV–outcome associations could state the sampling rate of each signal, report effects stratified by baseline variability where sample size allows, name the QRS detector and interpolation method, and show a QBA sensitivity band before presenting a dichotomised HRV analysis. Each step uses information already available to the analyst. The goal is not to establish a universal sampling-rate threshold — the appropriate rate depends on the HRV index and the variability range, as Zhou and colleagues and Burma and colleagues show [1,3] — but to make the influence of the chosen rate visible.

## 5. Conclusion

Differences in ECG sampling rate are a measurable source of bias in HRV–outcome research. Using sampling rate as a negative-control exposure and an empirically estimated distortion coefficient, we found a bounded effect on a continuous association and a larger effect on threshold-based classification. The method is a bias audit, not a causal design and not a clinical test. Reporting sampling rate, baseline-stratified effects and a quantitative sensitivity analysis would allow readers to judge how much of an HRV association survives the measurement process.

## Declarations

### Data availability

Code, aggregate statistics and figures are archived at Zenodo v2, DOI 10.5281/zenodo.24136999 (concept 10.5281/zenodo.24122064). Under the PhysioNet data-use agreement, waveforms and per-segment tables are not redistributed [35,37]. MIMIC-III, MIMIC-IV and PTB-XL were accessed as credentialed datasets [31–35].

### Ethics

The study was approved by the Ethics Committee of Shanghai Pudong New Area People’s Hospital (approval no. 2026-K82). The analysis used only de-identified, credentialed public datasets and involved no identifiable individual-level data; the PhysioNet data-use agreement terms governing derived aggregate results were observed.

### Author contributions and approval

All authors contributed to the study design and interpretation. All six authors read and approved the final manuscript; signed approval forms are retained by the corresponding author.

### Artificial-intelligence use disclosure

An AI language model assisted with English-language editing and with the verification of reference metadata. All scientific claims, numerical analyses and interpretations were authored and checked by the authors against the local analysis outputs.

## References

1. Zhou Y, Lindsey B, Snyder S, et al. Sampling rate requirement for accurate calculation of heart rate and its variability based on the electrocardiogram. Physiol Meas. 2024;45(2):025007. doi:10.1088/1361-6579/ad252d.

2. Hejjel L, Roth E. What is the adequate sampling interval of the ECG signal for heart rate variability analysis in the time domain? Physiol Meas. 2004;25(6):1405–1411. doi:10.1088/0967-3334/25/6/006.

3. Burma JS, Lapointe AP, Soroush A, et al. Insufficient sampling frequencies skew heart rate variability estimates. J Biomed Inform. 2021;123:103934. doi:10.1016/j.jbi.2021.103934.

4. Ellis R, Zhu B, Koenig J, Thayer JF, Wang Y. A careful look at ECG sampling frequency and R-peak interpolation on short-term measures of heart rate variability. Physiol Meas. 2015;36(9):1825–1842.

5. Béres S, Hejjel L. The minimal sampling frequency of the photoplethysmogram for accurate pulse rate variability parameters in healthy volunteers. Biomed Signal Process Control. 2021;68:102589. doi:10.1016/j.bspc.2021.102589.

6. Buchman TG, Stein PK, Goldstein B. Heart rate variability in critical illness and critical care. Curr Opin Crit Care. 2002;8(4):311–315. doi:10.1097/00075198-200208000-00007.

7. Ahmad S, Ramsay T, Huebsch L, et al. Continuous multi-parameter heart rate variability analysis heralds onset of sepsis in adults. PLoS One. 2009;4(8):e6642.

8. Fornasa F, et al. Heart rate variability as predictor of mortality in sepsis: a systematic review. PLoS One. 2018. PMC6133362.

9. Bento L, Fonseca-Pinto R, Póvoa P. Autonomic nervous system monitoring in intensive care as a prognostic tool: systematic review. Rev Bras Ter Intensiva. 2018;30(1):90–100. PMID 29340538.

10. Heart rate variability in critical care medicine: a systematic review. Intensive Care Med Exp. 2017. PMC5507939.

11. Lash TL, Fox MP, Fink AK. Applying Quantitative Bias Analysis to Epidemiologic Data. Springer; 2009. doi:10.1007/978-0-387-87959-8.

12. Fox MP, MacLehose RF, Lash TL. Applying Quantitative Bias Analysis to Epidemiologic Data. 2nd ed. Springer; 2021. doi:10.1007/978-3-030-82673-4.

13. Lash TL, Fox MP, MacLehose RF, et al. Good practices for quantitative bias analysis. Int J Epidemiol. 2014;43(6):1969–1985. doi:10.1093/ije/dyu149.

14. Greenland S. Basic methods for sensitivity analysis of biases. Int J Epidemiol. 1996;25(6):1107–1116. doi:10.1093/ije/25.6.1107.

15. Greenland S. Multiple-bias modelling for analysis of observational data. J R Stat Soc A. 2005;168(2):267–306. doi:10.1111/j.1467-985X.2004.00349.x.

16. Schneeweiss S. Sensitivity analysis and external adjustment for unmeasured confounders in epidemiologic database studies of therapeutics. Pharmacoepidemiol Drug Saf. 2006;15(5):291–303.

17. MacLehose RF, Kaufman S, Kaufman JS, Poole C. Bounding causal effects under uncontrolled confounding using counterfactuals. Epidemiology. 2005;16(4):548–555.

18. Quantitative bias analysis for mismeasured variables in health research: a review of software tools. BMC Med Res Methodol. 2025. doi:10.1186/s12874-025-02635-w.

19. Zeng Z, Wang J, Zuo H, Shu L. Quantifying systematic bias in observational causal estimates using negative controls: the bias attribution fraction. medRxiv. 2026. doi:10.64898/2026.10.08.26365019.

20. Lipsitch M, Tchetgen Tchetgen E, Cohen T. Negative controls: a tool for detecting confounding and bias in observational studies. Epidemiology. 2010;21(3):383–388. doi:10.1097/EDE.0b013e3181d61eeb.

21. Arnold BF, Ercumen A, Benjamin-Chung J, Colford JM. Negative controls to detect selection bias and measurement bias in epidemiologic studies. Epidemiology. 2016;27(5):637–641. doi:10.1097/EDE.0000000000000504.

22. Shi X, Miao W, Tchetgen Tchetgen EJ. A selective review of negative control methods in epidemiology. Curr Epidemiol Rep. 2020;7:119–137.

23. Tchetgen Tchetgen EJ. A general regression framework for a secondary outcome in case-control studies. Biostatistics. 2014;15(1):117–128. doi:10.1093/biostatistics/kxt041.

24. VanderWeele TJ, Ding P. Sensitivity analysis in observational research: introducing the E-value. Ann Intern Med. 2017;167(4):268–274. doi:10.7326/M16-2607.

25. Ding P, VanderWeele TJ. Sensitivity analysis without assumptions. Epidemiology. 2016;27(3):368–377. doi:10.1097/EDE.0000000000000457.

26. Schuemie MJ, Hripcsak G, Ryan PB, Madigan D, Suchard MA. Empirical confidence interval calibration for population-level effect estimation studies in observational healthcare data. PNAS. 2018;115(11):2571–2577.

27. Schuemie MJ, Ryan PB, DuMouchel W, Suchard MA, Madigan D. Interpreting observational studies: why empirical calibration is needed to correct p-values. Stat Med. 2014;33(2):209–218.

28. Hernán MA, Cole SR. Invited commentary: causal diagrams and measurement bias. Am J Epidemiol. 2009;170(8):959–962. doi:10.1093/aje/kwp293.

29. Hernán MA, Hernández-Díaz S, Robins JM. A structural approach to selection bias. Epidemiology. 2004;15(5):615–625.

30. Pirondini L, Diaz-Ordaz K, Palmer E, Keogh RH. Evaluating the impact of longitudinal treatment strategies in the presence of informative monitoring and time-dependent confounding. arXiv:2604.09898 [stat.AP]. 2026.

31. Johnson AEW, Bulgarelli L, Shen L, et al. MIMIC-IV, a freely accessible electronic health record dataset. Sci Data. 2023;10:1. doi:10.1038/s41597-022-01899-x.

32. Johnson AEW, Pollard TJ, Shen L, et al. MIMIC-III, a freely accessible critical care database. Sci Data. 2016;3:160035. doi:10.1038/s41597-016-0018-3.

33. Moody B, Moody G, Villarroel M, Clifford GD, Silva I. MIMIC-III Waveform Database (version 1.0). PhysioNet; 2020. doi:10.13026/c2607m.

34. Wagner P, Strodthoff N, Bousseljot RD, et al. PTB-XL, a large publicly available electrocardiography dataset. Sci Data. 2020;7:154. doi:10.1038/s41597-020-0495-6.

35. Goldberger AL, Amaral LAN, Glass L, et al. PhysioBank, PhysioToolkit, and PhysioNet: components of a new research resource for complex physiologic signals. Circulation. 2000;101(23):e215–e220.

36. Malik M, Bigger JT, Camm AJ, et al. Heart rate variability: standards of measurement, physiological interpretation, and clinical use. Circulation. 1996;93(5):1043–1065.

37. Pollard TJ, Moody BE, Lehman LH, et al. PhysioNet as a global platform for biomedical research. Nature Health. 2026;1(8):792–795. doi:10.1038/s44360-026-00096-z.

38. Carroll RJ, Ruppert D, Stefanski LA, Crainiceanu CM. Measurement Error in Nonlinear Models: A Modern Perspective. 2nd ed. Chapman & Hall/CRC; 2006.

39. Fuller WA. Measurement Error Models. Wiley; 1987.

40. Guo X, Lipsitz SR. Regression analysis with covariates that have heteroscedastic measurement error. Stat Med. 2007;26(19):3626–3643.

41. Hao Y. End-to-end waveform reanalysis of sampling-rate distortion in HRV (code and aggregates). Zenodo v2; 2026. doi:10.5281/zenodo.24136999(concept10.5281/zenodo.24122064).

42. Wang H, Zhang B, Lei Y, et al. From bias detection to distributional calibration: negative controls for shared systematic error in real-world evidence pipelines. medRxiv. 2026. doi:10.64898/2026.07.08.26357550.

43. Jayarathna T, Lui G, Breen P. Less is more: intelligent sparse sampling preserves R-peak detection and HRV accuracy down to 12 Hz. Zenodo v0.1-preprint. 2026. doi:10.5281/zenodo.20748922.

